# High SARS-CoV-2 attack rates following exposure during five singing events in the Netherlands, September-October 2020

**DOI:** 10.1101/2021.03.30.21253126

**Authors:** Anita A. Shah, Florien Dusseldorp, Irene K. Veldhuijzen, Margreet J.M. te Wierik, Alvin Bartels, Jack Schijven, Lucie C. Vermeulen, Mirjam J. Knol

## Abstract

**Background:** Previous reports suggest SARS-CoV-2 transmission risk increases during singing events. From September-October 2020, several clusters of COVID-19 cases among singing events were reported across the Netherlands. Our aim was to investigate whether singing increased SARS-CoV-2 transmission risk during these events.

**Methods:** Data from 5 events were retrospectively collected from spokespersons and singing group members via questionnaires. Information was consolidated with the National Notifiable Diseases Surveillance System. Specimens were requested for sequencing for point source and cluster assessment. We described outbreaks in terms of person, place and time and depicted potential SARS-CoV-2 transmission routes. A previously published model (AirCoV2) was used to estimate mean illness risk of 1 person through airborne transmission under various scenarios.

**Results:** Events included 9-21 persons (mean: 16), aged 20-89 years (median: 62). Response rates ranged 58-100%. Attack rates were 53-74%. Limited sequencing data was obtained from 2 events. Events lasted 60-150 minutes (singing: 20-120). Rooms ranged 320-3000m^3^. SARS-CoV-2 transmission likely occurred during all events; with a possible index case identified in 4 events. AirCoV2 showed 86% (54-100%) mean illness risk for 120 minutes of singing, smaller room (300m^3^), 1 air exchange/hour (ACH), and supershedder presence.

**Conclusions:** Droplet transmission and indirect contact probably caused some cases, but unlikely explain the high attack rates. AirCoV2 indicated that airborne transmission due to singing is possible in case of supershedder presence. Airflow expelling respiratory droplets >1.5m possibly influenced transmission. It is possible that singing itself increased SARS-CoV-2 transmission risk through airborne transmission.

**Summary:** This outbreak investigation among five singing events with high SARS-CoV-2 attack rates (53-74%) suggested that airflow expelling respiratory droplets >1.5m possibly influenced transmission and it is possible that singing itself increased SARS-CoV-2 transmission risk through airborne transmission.

## Introduction

Several outbreaks with high attack rates among singing groups, including several in the Netherlands, were described in literature and media from March until September 2020 [1–6] suggesting a possible elevated risk of severe acute respiratory syndrome coronavirus 2 (SARS-CoV-2) transmission for singing. It was unclear whether the outbreaks were the result of frequent and prolonged social contact (<1.5m) before, during, or after the singing event, or whether singing itself was a risk [7]. Singing groups stopped practicing from March 2020 in the Netherlands due to widespread SARS-CoV-2 transmission alongside other lockdown measures. Due to decreasing incidence of coronavirus disease 2019 (COVID-19) in the summer and easing of restrictions, group singing was allowed again in the Netherlands from July 2020. Specific recommendations for group singing included singing in zigzag formation and following ventilation advice guidelines, including using a room with mandatory ventilation rates for gatherings and regular venting when people were not in the room (e.g. during breaks) [7]. Face mask use in indoor places was not obligatory during this period. In the Netherlands, approximately one million singers participate in 24,000 choirs [8] and an estimated 70% of choirs resumed practicing from September 2020. From September through October 2020, there was a rapid increase in weekly COVID-19 incidence in the Netherlands from 31.4 to 391 per 100,000 [9].

From September through October 2020, we investigated clusters of COVID-19 cases among five singing events that were reported to the National Institute for Public Health and Environment (RIVM) in the Netherlands. Four reported clusters were related to choir rehearsals and one to a singing ensemble during a church service. An investigation was carried out to establish whether singing increased SARS-CoV-2 tranmission risk during these events. Here, we describe the outbreaks in terms of person, place and time and depict potential routes of SARS-CoV-2 transmission for each event.

## Methods

### Epidemiological investigations

Data on the five singing events were retrospectively first collected from a singing group spokesperson (organiser/singing group member) by phone or email and then an online questionnaire was sent to all singing group members. Questionnaire data was consolidated with the National Notifiable Diseases Surveillance System data, in order to deduce symptom onset date and positivity status. Formation diagrams were provided illustrating singing group member positions for each event. Diagrams were simplified and information aggregated to protect data confidentiality.

### Laboratory detection

During the investigation period, the testing policy in the Netherlands was that only persons experiencing COVID-19-like symptoms could be tested free of charge using reverse transcription polymerase chain reaction (RT-PCR) tests. However, four asymptomatic persons were also tested on their own accord. We followed up on laboratory specimens for all confirmed cases for whole SARS-CoV-2 genome sequencing for point source and cluster assessment.

### Definitions

An **outbreak confirmed case** was a person who was a singing group member (singer, conductor or musician) with a respiratory sample testing RT-PCR positive for SARS-CoV-2.

An **outbreak probable case** was a person who was a singing group member (singer, conductor or musician) and developed at least one of the following symptoms: cough, fever, sore throat, runny nose, increased or sudden loss of taste, loss of smell, fatigue, and shortness of breath, within 14 days following the singing event.

In one event, a preacher was considered a member of the singing group due to being in close proximity of singing group members.

**Droplet transmission** is infection spread through exposure to virus-containing respiratory droplets (i.e., larger and smaller droplets and particles) exhaled by an infectious person [10]. Transmission is most likely to occur when someone is close to the infectious person, generally within 1.5m distance (e.g. contact with respiratory droplets after a cough). Respiratory droplets can be expelled further than 1.5m because of air currents or forceful ejections (e.g. violent sneeze) [11, 12].

**Indirect contact transmission** is infection after contact with an article or surface that has become contaminated [10].

**Airborne transmission** is infection spread through exposure to those virus-containing respiratory droplets comprised of smaller droplets and particles that can remain suspended in the air and, therefore be transported over longer distances (several metres) and time (typically hours) [10].

### Aerosol transmission model

We used model AirCoV2 (version 1.5) described by Schijven et al. [13] to assess under which circumstances aerosols production by singing could have led to attack rates observed in the five singing events. The developed scenarios are based on event circumstances, although insufficient information was available to exactly simulate the circumstances. The model assumes an even distribution of aerosols containing virus particles across the space, and relatively high estimates of virus infectivity [13]. The model was applied for 20 scenarios encompassing a small and large room (300 vs 3000m^3^), exposure times of 30, 60 and 120 minutes, no ventilation, one or six air exchanges per hour (ACH). The model also includes concentration of 10^7^ or 10^10^ virus RNA copies per mL mucus (where 10^10^ represents supershedder presence) [13, 14].

## Results

### Singing event descriptions

We report on five singing events across the Netherlands with minimum attack rates (including confirmed and probable cases) from 53–74% (Table 1). Online questionnaire response rates were 58–100%. Events included 9–21 persons, aged 20–89 years (Table 1). In all events, transmission likely occurred during the event itself as this was the only common place and time that all affected singing group members were together. Cases were widely dispersed throughout the room in all events. From event room size descriptions, all events except event 2, had adequate space for members to keep 1.5m distance from eachother (Table 1). In all events, except event 3, members stated that they kept 1.5m distance from eachother (Table 2). In events 1, 3, and 4, members reported feeling an air draft, however specific airflow information is not known (Table 2). Ventilation and other mechanisms used in each event is specified in Table 1.

**Table 1.** Characteristics for each singing event and their respective venues from September–October 2020.

|  | Singing event 1 | Singing event 2 | Singing event 3 | Singing event 4 | Singing event 5 |
| --- | --- | --- | --- | --- | --- |
| **Characteristics of singing group members** |  |  |  |  |  |
| Singing group members | 19 | 21 | 15 | 14 | 9 |
| Case definitions |  |  |  |  |  |
| Confirmed | 14 | 13 | 8 | 7 | 6 |
| Probable | 0 | 1 | 0 | 1 | 0 |
| Attack rate** | 74%<br>(14/19)* | 67%<br>(14/21) | 53%<br>(8/15) | 57%<br>(8/14) | 67%<br>(6/9)* |
| Questionnaire response rate | 58%<br>(11/19) | 95%<br>(20/21) | 73%<br>(11/15) | 100%<br>(14/14) | 78%<br>(7/9) |
| Sex |  |  |  |  |  |
| Female | 11* | 12 | 7 | 8 | 6* |
| Male | 6* | 8 | 4 | 6 | 1* |
| Median age (range) | 74<br>(60-89)* | 62.5<br>(54-74) | 51<br>(32-70) | 57.5<br>(32-74) | 41<br>(20-55)* |
| Specimens sequenced | 0 | 0 | 0 | 2 | 5 |
| Previously tested for SARS-CoV-2 since January 2020 |  |  |  |  |  |
| Positive | 0 | 0 | 0 | 0 | 0 |
| Negative | 0 | 2 | 3 | 3 | 1 |
| Not tested | 10 | 18 | 8 | 11 | 6 |
| Missing | 1 | 0 | 0 | 0 | 0 |
| **Characteristics of venue where singing event took place** |  |  |  |  |  |
| Venue type | Hall | Hall | Hall | Hall | Church |
| Size of venue (l x w x h)m<sup>3</sup> | 510<br>(14x14x2.6) | 80m<sup>2</sup><br>(+ ~8m roof) | 561<br>(11x8.5x6) | ~320 | 3000<br>(20x15x10) |
| Duration of event (mins) | 90 | 120 | 150 | 120 | 60 |
| Duration of singing (mins) | 50 | ~80 | 120 | ~90 | 20 |
| Duration of break (mins) | 15 | 5 | 15 | 15 | NA |
| Natural ventilation |  |  |  |  |  |
| Number of doors open |  |  |  |  |  |
| Facing inside | 1 | 2 | 1 | 2 | 2 |
| Facing outside | 1 | 1 | 0 | 0 | 0 |
| Number of windows open | 0 | 2 | 2 | 6 | 1 |
| Additional ventilation | None | None | Ceiling ventilation | Possible mechanical ventilation | None |
| Other mechanism | Heat exchanger | None | None | None | Air heating |

\*Data reported here is combined from the National Notifiable Surveillance Disease System and questionnaire responses.
\*\*Attack rate includes confirmed and probable cases.

**Table 2.** Potential exposures within and outside of the singing events for confirmed and probable COVID-19 cases among singing groups from September–October 2020.

|  |  | Singing<br>event 1<br>(n = 8) | Singing<br>event 2<br>(n = 14) | Singing<br>event 3<br>(n = 8) | Singing<br>event 4<br>(n = 8) | Singing<br>event 5<br>(n = 6) |
| --- | --- | --- | --- | --- | --- | --- |
| Kept 1.5m distance during the rehearsal/ performance | Yes | 8 | 14 | 7 | 8 | 6 |
|  | No | 0 | 0 | 1 | 0 | 0 |
| Kept 1.5m distance during the break | Yes | ? | ? | 6 | 8 | NA |
|  | No | ? | ? | 2 | 0 | NA |
| Had contact before/after the rehearsal/ performance | Yes | 2 | 2 | 3 | 2 | 2 |
|  | No | 6 | 12 | 5 | 6 | 4 |
| Kept 1.5m distance before/ after the singing event | Yes | 2 | 2 | 0 | 0 | 2 |
|  | No | 0 | 0 | 3 | 2 | 0 |
| Travelled together to/from the singing event | Yes | 3 | 3 | 2 | 5 | 0 |
|  | No | 5 | 11 | 6 | 3 | 6 |
| Kept 1.5m distance during travel | Yes | 0 | 0 | 0 | 1 | NA |
|  | No | 3 | 3 | 1 | 4 | NA |
| Felt (cold) airflow during the singing event | Yes | 2* | ?* | 4 | 2 | 1 |
|  | No | ?* | ?* | 4 | 6 | 5 |
| Toilet used during the break | Yes | ? | ? | 6 | 1 | 4 |
|  | No | ? | ? | 2 | 7 | 2 |
| Sang in another singing group 14 days prior to singing event | Yes | 1 | 1 | 0 | 0 | 1 |
|  | No | 7 | 13 | 8 | 8 | 5 |
| Had contact with person tested positive for SARS-CoV-2 in the 14 days prior to singing event | Yes | 0 | 0 | 1 | 0 | 1 |
|  | No | 8 | 14 | 7 | 8 | 5 |
| Had contact with person tested positive for SARS-CoV-2 in the 14 days following the singing event | Yes | 2 | 1 | 0 | 0 | 1 |
|  | No | 6 | 13 | 8 | 8 | 5 |
| Had contact with person tested positive for SARS-CoV-2 (date not specified) | Yes | 1 | 0 | 1 | 0 | 0 |

\* Information for these two singing groups was extracted from comments in the singing group member questionnaire. Question investigating airflow was later added to the singing group member questionnaire following spontaneous reporting.

### Singing event 1

In singing event 1, 14 confirmed cases were identified out of 19 singing group members. Twelve of 14 confirmed cases developed symptoms within 11 days of the event (Figure 1). One confirmed case was hospitalised. A single index case could not be clearly identified as seven persons had their symptom onset three days following the event (Figure 1). Three pairs travelled to and from the event together by car. In one pair, one person may have infected the other outside of the event as there were eight days between their symptom onset and they lived together. In the other two pairs, only one person in each pair tested positive. During the break, movement was limited as members remained in place and staff served coffee. Additional information was not available for staff. Event room doors were kept open and thus, touching of door handles was limited.

**Figure 1.**
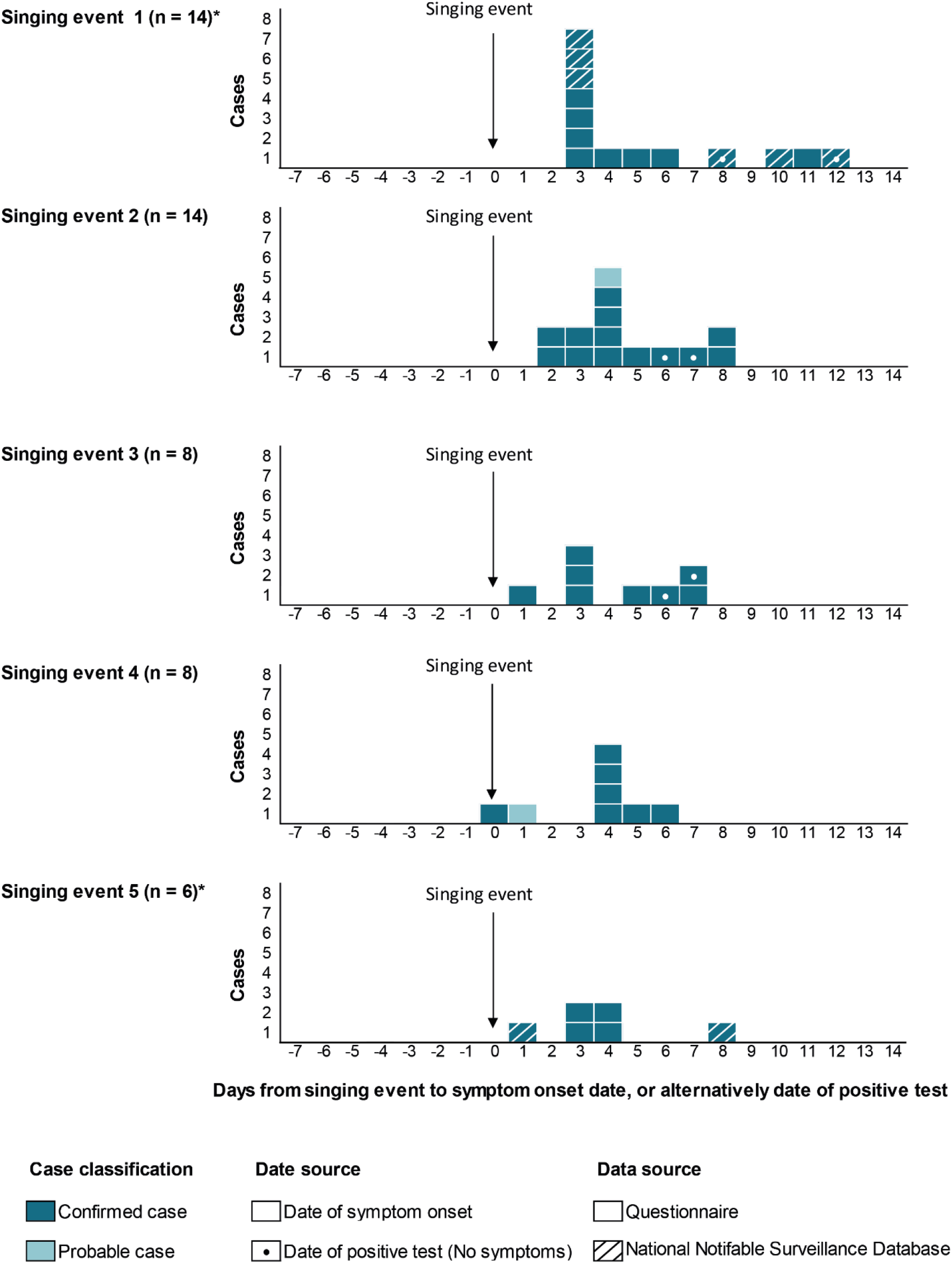
Confirmed and probable COVID-19 cases in each singing event, September– October 2020 by date of symptom onset, or alternatively by date of positive test. *Data is combined from the National Notifiable Surveillance Disease System and questionnaire responses.

### Singing event 2

In singing event 2, 13 confirmed cases were identified out of 21 singing group members. Eleven of 13 confirmed cases developed symptoms within 7 days of the event (Figure 1). One probable case was also reported with symptom onset 4 days following the event. Two possible index cases with symptom onset two days following the event were identified (Figure 1). Six members travelled together by bicycle, and two pairs by car. Of the six who travelled by bicycle, two were confirmed cases. Among the two pairs who travelled by car; one pair included two confirmed cases and the other pair included one confirmed case. Members used individual sheet music and available toilets were spacious. Touchable shared surfaces were limited.

### Singing event 3

In singing event 3, eight confirmed cases were identified out of 15 singing group members. Six of eight confirmed cases experienced symptoms within 7 days of the event (Figure 1). A possible index case was identified with symptom onset on the day after the event (Figure 1). This case also mentioned contact with a positive case, however, date was not specified. An additional person reported contact with a positive case in the 14 days prior to event however, this person’s symptom onset was seven days following the event, therefore they are unlikely to be an index case (Table 2). Two pairs travelled together by car, and both tested positive in one pair, whereas the other pair included one person who tested negative and one person who was not tested. Common surfaces were limited with spacious separate toilets by gender.

### Singing event 4

In singing event 4, seven confirmed cases were identified out of 14 singing group members. All developed symptoms within six days of the event (Figure 1). One probable case was also reported with symptom onset 1 day following the event. A possible index case was identified who had symptom onset on the day of event (Figure 1). Six singing group members travelled together; two pairs by bicycle (all tested positive) and one pair by car (one tested positive, and the other was not tested, nor had symptoms). During the break, a coffee machine was used which required pushing a button. Three to four members assisted stacking chairs. Available sequencing revealed two identical strains in two persons positioned on opposite sides of the room (Table 1).

### Singing event 5

In singing event 5, six confirmed cases were identified out of 9 singing group members. All developed symptoms within eight days of the event (Figure 1). A possible index case was identified with symptom onset on the day after the event (Figure 1). This case was likely to have had contact with confirmed cases during work in the week prior to event. No common objects were reported to have been touched and individual microphones were used throughout the event. Sequencing revealed four out of five identical strains in persons positioned near each other. Screens were placed in front of singing group members and between church service attendees. It is not known how screen use influenced transmission. Data was available for 58% of church service attendees (morning and afternoon). A maximum of 30 attendees were allowed at each service and singing group members were only present in the afternoon. Only one positive case was known among the attendees with possible exposure at the afternoon service. It is unclear whether this case is linked to confirmed cases among the singing group members as sequencing was not done.

### Aerosol transmission model

The AirCoV2 model showed that the mean risk of illness of one person was 86% (54-100%) in a smaller room (300m^3^), 120 minutes of exposure time, one ACH, and with the presence of a supershedder (Table 3, Scenario 8). In a 10-fold larger room (3000m^3^), the mean risk of illness of one person was approximately four fold lower (24%; Table 3, Scenario 18). Event room sizes ranged from 320-3000m^3^. Halving the exposure time (60 minutes), reduced the mean risk of illness by one-third (58%; Table 3, Scenario 5). Singing events ranged from 60-150 minutes with singing duration from 20-120 minutes. Also increasing the ventilation to six ACH (9L/sec/person) reduced the dose and risk by approximately one-third (54%; Table 3, Scenario 9). Exact ACH for each event was not known. Based on received information on ventilation measures, events 1 and 5 could have had three ACH or more. For other events, one ACH or less is more likely. The mean risk of illness in case of supershedder presence was 94% with no ACH, exposure time of 120 minutes, in a small room (Table 3, Scenario 7) compared to 0.48% with no supershedder present (Table 3, Scenario 10). In the AirCoV2 model, the mean probability of illness fell within the range or was higher than the observed attack rates for singing events described in this outbreak investigation in scenarios with at least 60 minutes of singing in a small room, and in scenarios with 120 minutes of singing in a large room with no or little ventilation, with supershedder presence. Overall, the model indicated that high virus concentrations, i.e. supershedder presence, are required to explain high attack rates via this transmission route.

**Table 3.** Dose and illness risks from simulations with AirCoV2 version 1.5.

|  | m<sup>3</sup> | Q | T | C | Cumulative dose (virus RNA copies)<br>/person |  |  |  | Illness risk/person/event |  |  |  |
| --- | --- | --- | --- | --- | --- | --- | --- | --- | --- | --- | --- | --- |
|  |  |  |  |  | Mean | 5% | 50% | 95% | Mean | 5% | 50% | 95% |
| 1 | 300 | 0 | 30 | 10 | 540 | 140 | 430 | 1300 | 29% | 9.4% | 26% | 60% |
| 2 | 300 | 1 | 30 | 10 | 470 | 120 | 370 | 1100 | 26% | 8.1% | 23% | 54% |
| 3 | 300 | 6 | 30 | 10 | 250 | 66 | 200 | 620 | 16% | 4.5% | 13% | 35% |
| 4 | 300 | 0 | 60 | 10 | 2000 | 530 | 1600 | 4800 | 66% | 31% | 67% | 97% |
| 5 | 300 | 1 | 60 | 10 | 1500 | 390 | 1200 | 3700 | 58% | 24% | 57% | 92% |
| 6 | 300 | 6 | 60 | 10 | 610 | 160 | 490 | 1500 | 32% | 11% | 29% | 64% |
| 7 | 300 | 0 | 120 | 10 | 6900 | 1800 | 5500 | 17000 | 94% | 72% | 98% | 100% |
| 8 | 300 | 1 | 120 | 10 | 4300 | 1100 | 3400 | 10000 | 86% | 54% | 91% | 100% |
| 9 | 300 | 6 | 120 | 10 | 1300 | 350 | 1100 | 3200 | 54% | 22% | 53% | 90% |
| 10 | 300 | 0 | 120 | 7 | 6.9 | 1 | 5 | 18 | 0.48% | 0.07% | 0.35% | 1.3% |
| 11 | 3000 | 0 | 30 | 10 | 55 | 13 | 43 | 130 | 3.7% | 0.91% | 3% | 8.9% |
| 12 | 3000 | 1 | 30 | 10 | 47 | 11 | 37 | 110 | 3.2% | 0.77% | 2.6% | 7.6% |
| 13 | 3000 | 6 | 30 | 10 | 26 | 6 | 20 | 62 | 1.8% | 0.42% | 1.4% | 4.2% |
| 14 | 3000 | 0 | 60 | 10 | 200 | 52 | 160 | 490 | 13% | 3.6% | 11% | 29% |
| 15 | 3000 | 1 | 60 | 10 | 150 | 38 | 120 | 370 | 9.9% | 2.6% | 8.1% | 23% |
| 16 | 3000 | 6 | 60 | 10 | 62 | 15 | 49 | 150 | 4.2% | 1% | 3.4% | 10% |
| 17 | 3000 | 0 | 120 | 10 | 700 | 180 | 560 | 1700 | 35% | 12% | 32% | 70% |
| 18 | 3000 | 1 | 120 | 10 | 430 | 110 | 340 | 1000 | 24% | 7.5% | 21% | 52% |
| 19 | 3000 | 6 | 120 | 10 | 130 | 34 | 110 | 330 | 8.8% | 2.4% | 7.2% | 20% |
| 20 | 3000 | 0 | 120 | 7 | 0.7 | 0 | 0 | 3 | 0.049% | 0% | 0% | 0.21% |

m<sup>3</sup>: room size, small room 10x10x3=300m<sup>3</sup>, large room 20x25x6=3000 m<sup>3</sup>;
Q: air exchanges/hour; T: exposure time (minutes); C: virus RNA copies/mL, 10=10<sup>10</sup> and 7=10<sup>7</sup>.
Scenarios:
- Always one person as source, 15 persons exposed.
- Fraction of virus RNA copies infectious to cell culture: 0.0125 (1/80).
- Fraction of those viruses leading to illness: 0.056 (1/18).
- So, fraction of virus RNA copies leading to illness: 0.0007 (1/1440).
- Probability of at least 10<sup>7</sup> RNA copies/ml in mucus is 66%.
- Probability of at least 10<sup>8</sup> RNA copies/ml in mucus is 36%.

## Discussion

Events had high attack rates (53-74%) demonstrating the high risk in these settings. Based on available epidemiological information, it is likely that SARS-CoV-2 transmission for most singing group members occurred at the singing events itself except those living together. In these five events, cases had little to no contact outside of events except for those who lived and travelled together to the event. At least six persons lived together and all six were confirmed cases. In all events, at least one singing group member reported their symptom onset between 0–3 days following the event (Figure 1). These singing group members were potentially contagious during the events and this corresponds with known SARS-CoV-2 transmission dynamics with higher levels of shedding just prior to development of symptoms [15, 16]. A possible index case could be identified in four out of five clusters according to available information.

SARS-CoV-2 transmission routes include droplet and indirect contact transmission [10, 17]. Dutch national advice for singing groups as well as in other countries is directed at preventing these transmission routes [7, 18]. In general, the five singing groups tried to adhere to this advice. From available information, droplet transmission through prolonged social contact (within 1.5m) during the event itself seemed unlikely. Air currents from person to person because of open windows and doors or mechanical ventilation may have increased SARS-CoV-2 droplet transmission during these events, as droplets could have moved over longer distances (>1.5m) [19]. It remains possible that even if there is adequate ventilation but air currents are present, the increased dispersion of droplets produced by singing may be sufficient to increase droplet transmission. Although there is insufficient evidence regarding air currents, this possibility cannot be ruled out. ECDC guidelines recommend that direct airflow should be diverted away from groups of individuals to avoid pathogen dispersion from infected subjects and transmission [20] and Dutch national guidelines recommend airing out indoor spaces (eg. leaving doors/windows opposite eachother wide open for 10-15 minutes) to create a draft during breaks [21].

Previous studies have shown SARS-CoV-2 transmission risk through indirect contact transmission to be low [22, 23]. Limited shared surfaces were present and only several singing group members reported touching common surfaces, therefore there is a low likelihood that this transmission route contributed to the high attack rates observed.

Singing expels approximately 10–15 times as much aerosols as speaking [13]. Additionally, singing increases aerosol dispersion compared to speaking and the amount of aerosols expelled increases with voice loudness [13, 24–27]. ACH was not obtained and event descriptions did not provide enough information to determine its role. Event rooms were generally large in size. If the air was not fully mixed, it may have been possible that air in the exhaled plume of the infectious person would have higher viral concentrations than the air in the far corner of the room, and thus gave rise to higher exposure closer to the infectious person. Airborne transmission is a possible route of transmission and has been described previously in a similar context [28]. In all events, cases were dispersed throughout the room which may be consistent with airborne transmission. According to the AirCov2 model, supershedder presence in the room is needed to achieve relevant risks of illness by aerosol transmission [19, 29–31].

From September to October 2020, 1.4% persons were infected in the Dutch population. Given that 2.7% of infected persons may have been a possible supershedder (10^10^ virus mL in mucus) [13], then an estimated 95 supershedders of 3528 contagious persons may have been present among 252,000 singing group members (~15 singers in each singing group) from September to October 2020. Therefore, supershedder presence among the five events is theoretically possible, however we cannot confirm supershedder presence since CT-values were not available for confirmed cases. Additionally, multiple source cases may have been present in at least two events which could have contributed to the high attack rates.

Our study has several limitations. Due to the observational nature of this study, it was difficult to reconstruct exact circumstances of the clusters and proportions attributable to each possible transmission route could not be deduced. Secondly, there was a low number of specimens for phylogenetic analysis due to limited specimen storage time (usually one week) during the investigation period. Also, a large number of laboratories were involved coupled with high work load for the public health services. Furthermore, asymptomatic cases may have been missed as not all persons were tested. Therefore, the number of possible source cases present at the events could not be identified in order to confirm whether singing group members were infected by a common source. Additionally, exact circumstances regarding airflow direction for each event were not known, and could not be accounted for in the model. Airflow was also dependent on weather for events using natural ventilation. Lastly, due to the retrospective nature of the questionnaires, recall bias could have affected the participants’ responses indicating the likeliness of different transmission routes. However, almost all members described similar circumstances under which events occurred.

## Conclusions

These outbreaks with high attack rates demonstrate the potential for SARS-CoV-2 transmission linked to singing events. In conclusion, our findings suggest that the described outbreaks were probably caused by a combination of different transmission routes as none of the possible transmission routes could be ruled out. Indirect contact and droplet transmission (<1.5m) may have occurred and may have been the cause of some cases, but it is unlikely to explain the high attack rates. The described AirCoV2 model indicated that airborne transmission (via infectious droplets/ aerosols over longer distances) was possible if a supershedder was present. Additionally, multiple index cases may have been present. However, the airflow as reported may also have expelled respiratory droplets over longer distances and previous studies have shown that directional airflow may possibly influence transmission [12, 32].

Further research is needed into the role of airflow and SARS-CoV-2 transmission dynamics in singing groups. Additionally, increased phylogenetic analysis should be performed to identify potential source cases to better assess clusters. Serology could also be performed to identify susceptible cases. In the clusters described here, it is possible that singing itself increased SARS-CoV-2 transmission risk through airborne transmission. As COVID-19 measures are eased and group gatherings are allowed, specific recommendations regarding group singing may be needed, although with increasing vaccination coverage these may be loosened again.

## Data Availability

The datasets generated during and/or analysed during the current study are not publicly available but are available from the corresponding author on reasonable request.

## Authors’ contributions

All authors contributed to the outbreak investigations described in this report. AS wrote the first draft of the manuscript and coordinated the revision of the manuscript. JS and LV contributed to the Aerosol transmission modelling outputs. All authors contributed to the interpretation of the data, critically revised the manuscript and approved the final version.

## Acknowledgements

We are thankful to staff at Municipal Health Services, laboratories and clinicians for reporting and investigating COVID-19 cases. We gratefully acknowledge the affected choir focal points for their trustful sharing of information and this study was only possible thanks to the collaboration of affected individuals from the outbreaks. We thank Harry Vennema for carrying out phylogenetic analysis and Atze Boerstra for ventilation advice during this outbreak investigation.

## Ethical statement

The outbreak investigations were carried out in accordance with the Dutch Public Health Act. Participation in the questionnaire was voluntary and online informed consent was obtained.

## References

1. Hamner, L., et al., High SARS-CoV-2 Attack Rate Following Exposure at a Choir Practice - Skagit County, Washington, March 2020. MMWR Morb Mortal Wkly Rep, 2020. 69(19): p. 606–610.

2. Gelderland, O. Koor Heerde verloor zes leden aan corona: 40 procent werd ziek. 2020 20 November 2020]; Available from: https://www.omroepgelderland.nl/nieuws/2448308/Koor-Heerde-verloor-zes-leden-aan-corona-40-procent-werd-ziek.

3. Gelderlander, D. Ook veel ‘coronagevallen’ bij vrouwenkoor Between Two rivers dat in Elst repeteerde. 2020 20 November 2020]; Available from: https://www.gelderlander.nl/overbetuwe/ook-veel-coronagevallen-bij-vrouwenkoor-between-two-rivers-dat-in-elst-repeteerde~ac080406/.

4. Oost, R. Corona in Hasselt Musicalkoor extra zwaar getroffen: dirigent ging door oog van de naald. 2020 [cited 2020; Available from: https://www.rtvoost.nl/nieuws/328138/Corona-in-Hasselt-Musicalkoor-extra-zwaar-getroffen-dirigent-ging-door-oog-van-de-naald.

5. Times, T. Did singing together spread COVID-19? 2020 20 November 2020]; Available from: https://www.taipeitimes.com/News/world/archives/2020/05/18/2003736628.

6. Limburger, D. Hoe een gouden jaar een zwart jaar werd: zangkoren recht in het hart getroffen door corona. 2020 20 November 2020]; Available from: https://www.limburger.nl/cnt/dmf20200517_00160696/hoe-een-gouden-jaar-een-zwart-jaar-werd-voor-zangkoren-gronsveld-en-eijsden.

7. RIVM. Koren en zangensembles. 2020 1 July 2020 20 November 2020]; Available from: https://lci.rivm.nl/koren-zangensembles.

8. Association, E.C. Singing Europe. 2015 20 November 2020]; Available from: https://europeanchoralassociation.org/wp-content/uploads/2019/01/singingeurope_report.pdf.

9. RIVM. Wekelijkse update epidemiologische situatie COVID-19 in Nederland. 2020 20 November 2020]; Available from: https://www.rivm.nl/coronavirus-covid-19/actueel/wekelijkse-update-epidemiologische-situatie-covid-19-in-nederland.

10. CDC. Scientific Brief: SARS-CoV-2 and Potential Airborne Transmission. 2020 5 October 2020 [cited 2021 25 January]; Available from: https://www.cdc.gov/coronavirus/2019-ncov/more/scientific-brief-sars-cov-2.html.

11. Bourouiba, L., Turbulent Gas Clouds and Respiratory Pathogen Emissions: Potential Implications for Reducing Transmission of COVID-19. Jama, 2020. 323(18): p. 1837–1838.

12. Kwon, K.S., et al., Evidence of Long-Distance Droplet Transmission of SARS-CoV-2 by Direct Air Flow in a Restaurant in Korea. J Korean Med Sci, 2020. 35(46): p. e415.

13. Schijven, J.F., et al., Quantitative risk assessment for airborne transmission of SARS-CoV-2 via breathing, speaking, singing, coughing and sneezing Environmental Health Perspectives, submitted. medRxiv, 2020: p. 2020.07.02.20144832.

14. Kleiboeker, S., et al., SARS-CoV-2 viral load assessment in respiratory samples. J Clin Virol, 2020. 129: p. 104439.

15. McAloon, C., et al., Incubation period of COVID-19: a rapid systematic review and meta-analysis of observational research. BMJ Open, 2020. 10(8): p. e039652.

16. Xie, Y., et al., Epidemiologic, clinical, and laboratory findings of the COVID-19 in the current pandemic: systematic review and meta-analysis. BMC Infectious Diseases, 2020. 20(1): p. 640.

17. WHO. Transmission of SARS-CoV-2: implications for infection prevention precautions. 2020 [cited 2021 17 February]; Available from: https://www.who.int/news-room/commentaries/detail/transmission-of-sars-cov-2-implications-for-infection-prevention-precautions.

18. England, P.H. COVID-19: suggested principles of safer singing. 2020 [cited 2021 17 February]; Available from: https://www.gov.uk/government/publications/covid-19-suggested-principles-of-safer-singing/covid-19-suggested-principles-of-safer-singing.

19. Bahl, P., et al., Droplets and Aerosols Generated by Singing and the Risk of Coronavirus Disease 2019 for Choirs. Clinical Infectious Diseases, 2020.

20. ECDC. Heating, ventilation and air-conditioning systems in the context of COVID-19: first update. 2020 20 November 2020]; Available from: https://www.ecdc.europa.eu/en/publications-data/heating-ventilation-air-conditioning-systems-covid-19.

21. RIVM. Ventilatie en COVID-19. 2020 20 November 2020]; Available from: https://lci.rivm.nl/ventilatie-en-covid-19.

22. Pitol, A.K. and T.R. Julian, Community Transmission of SARS-CoV-2 by Surfaces: Risks and Risk Reduction Strategies. Environmental Science & Technology Letters, 2021. 8(3): p. 263–269.

23. CDC. Science Brief: SARS-CoV-2 and Surface (Fomite) Transmission for Indoor Community Environments. 2021 [cited 2021 10 June]; Available from: https://www.cdc.gov/coronavirus/2019-ncov/more/science-and-research/surface-transmission.html#ref8.

24. Echternach, M., et al., Impulse Dispersion of Aerosols During Singing and Speaking: A Potential COVID-19 Transmission Pathway. Am J Respir Crit Care Med, 2020.

25. Asadi, S., et al., Aerosol emission and superemission during human speech increase with voice loudness. Sci Rep, 2019. 9(1): p. 2348.

26. Gregson, et al., Comparing the Respirable Aerosol Concentrations and Particle Size Distributions Generated by Singing, Speaking and Breathing. 2020.

27. Mürbe D K.M., Lange J, Rotheudt H, Fleischer M., Aerosol emission is increased in professional singing. OSF Preprints, 2020.

28. Katelaris, A.L., et al., Epidemiologic Evidence for Airborne Transmission of SARS-CoV-2 during Church Singing, Australia, 2020. Emerg Infect Dis, 2021. 27(6): p. 1677–1680.

29. Buonanno, G., L. Morawska, and L. Stabile, Quantitative assessment of the risk of airborne transmission of SARS-CoV-2 infection: prospective and retrospective applications. medRxiv, 2020: p. 2020.06.01.20118984.

30. Chen, P.Z., et al., Heterogeneity in transmissibility and shedding SARS-CoV-2 via droplets and aerosols. medRxiv, 2020: p. 2020.10.13.20212233.

31. Miller, S.L., et al., Transmission of SARS-CoV-2 by inhalation of respiratory aerosol in the Skagit Valley Chorale superspreading event. Indoor Air, 2020.

32. Lu, J., et al., COVID-19 Outbreak Associated with Air Conditioning in Restaurant, Guangzhou, China, 2020. Emerg Infect Dis, 2020. 26(7): p. 1628–1631.

