## Supplementary Figure 1 for "High SARS-CoV-2 attack rates following exposure during five singing events in the Netherlands, September-October 2020"

**Supplentary Figure 1. Singing group members in each singing event, September–October 2020 by COVID-19 test result.***

+ Tested positive
− Tested negative
⊗ Not tested
? No response

**a. Singing event 1 (n = 19)****

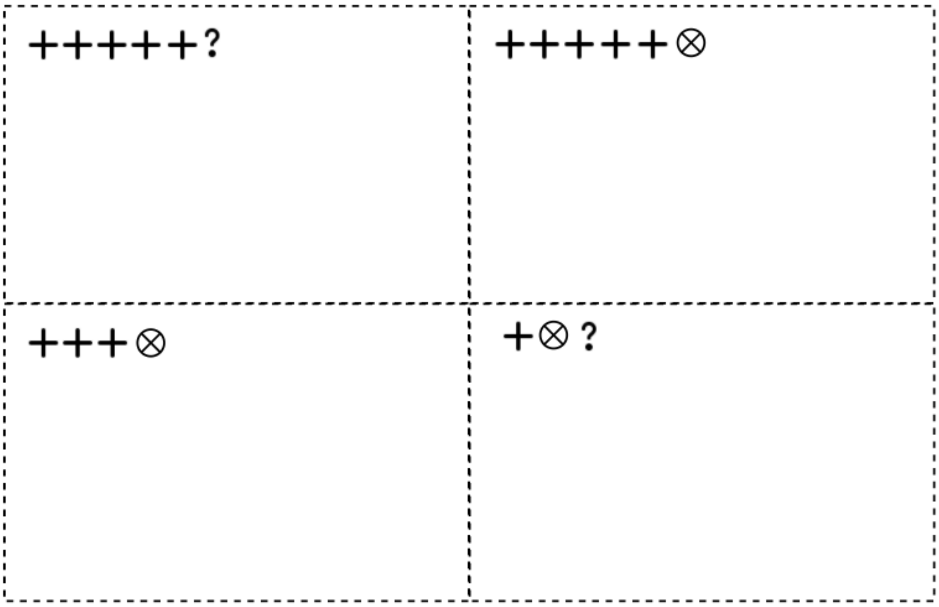


**b. Singing event 2 (n=21)**

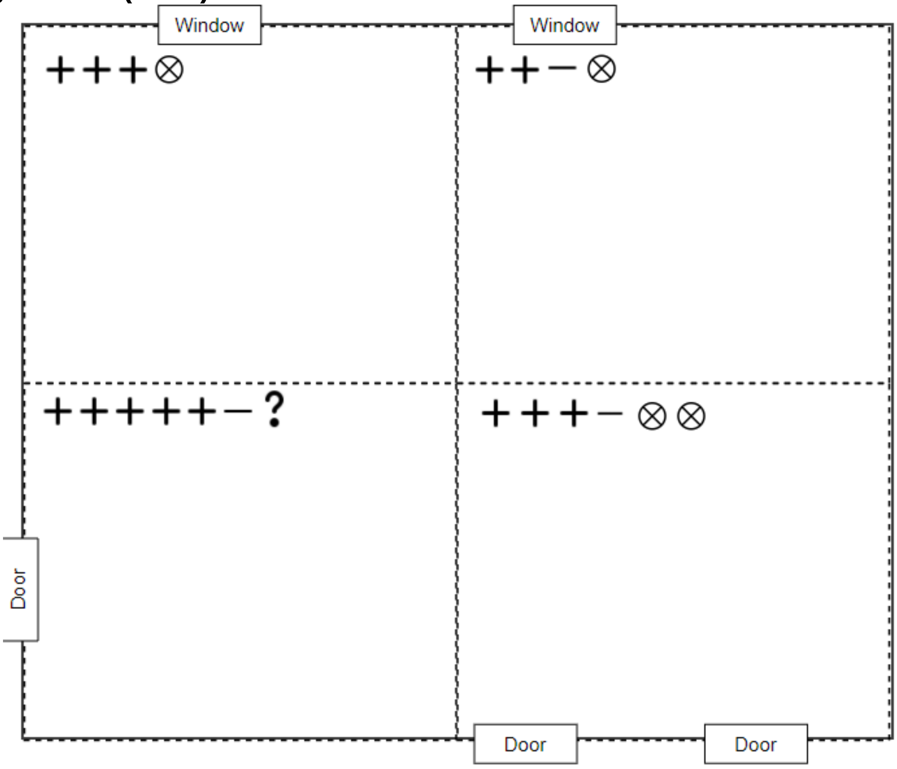

c. **Singing event 3 (n = 15)**

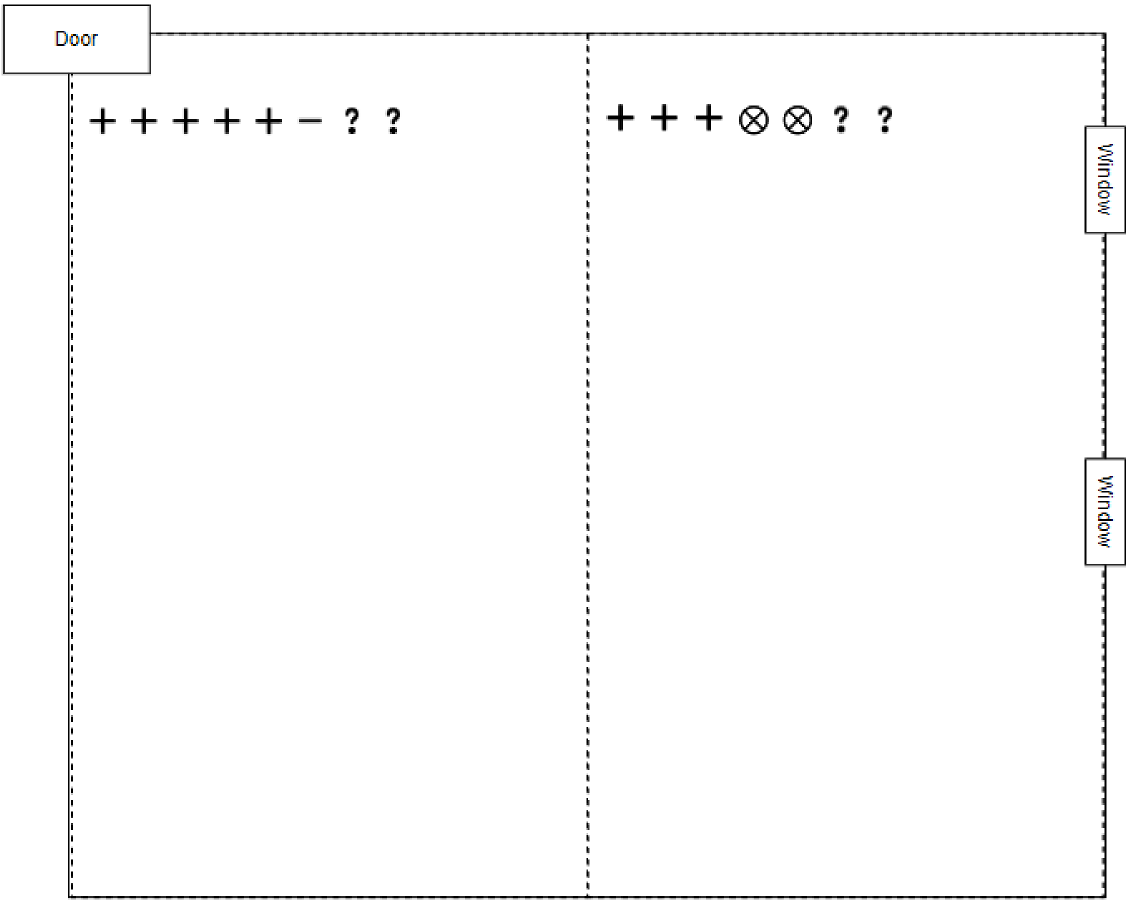


d. **Singing event 4 (n=14)**

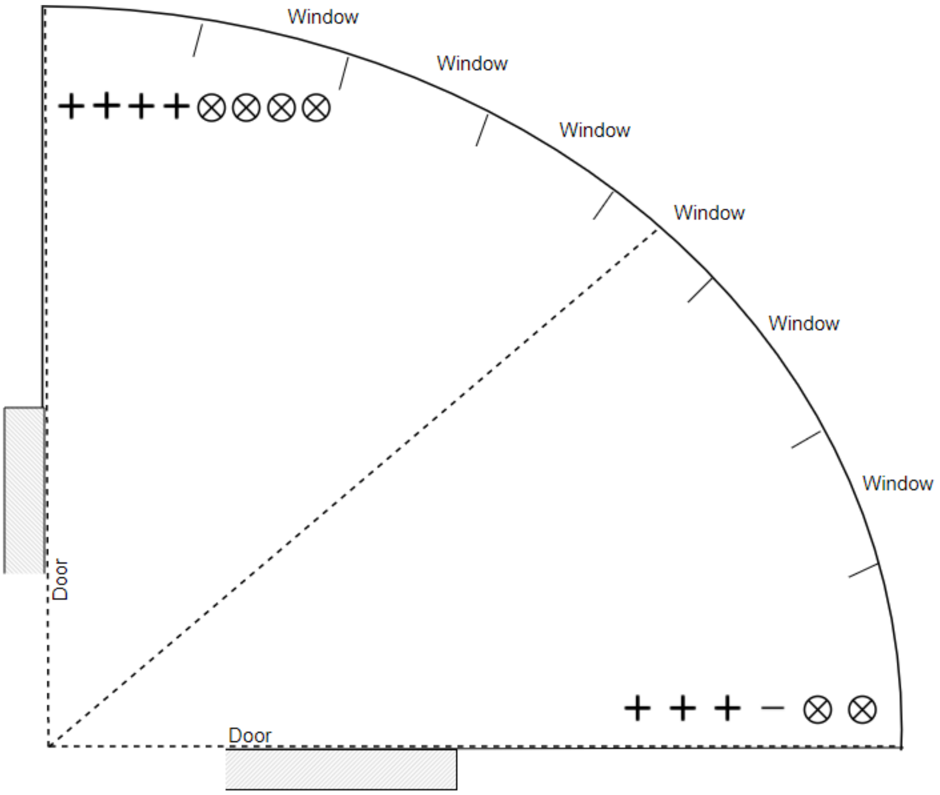

### e. Singing event 5 (n = 9)

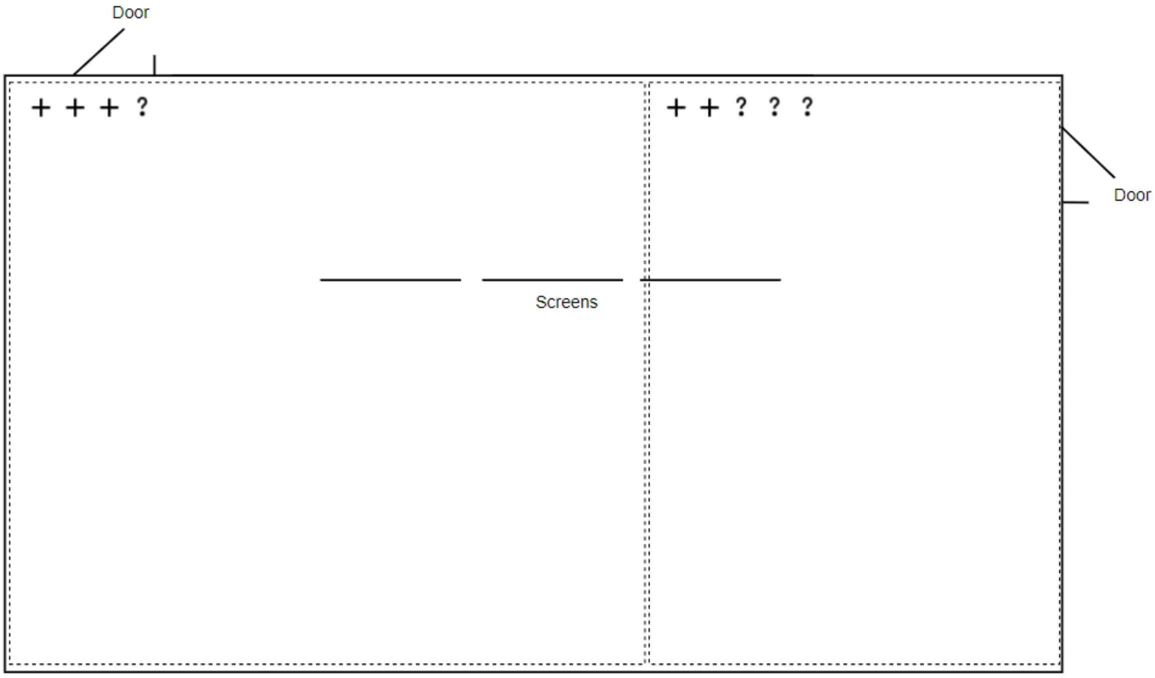


*Formation diagrams may not be an accurate representation of the room dimensions. Additionally, diagrams have been simplified and information was aggregated to protect data confidentiality.

**Exact placement of doors and windows is unknown.
